# The development of a multidisciplinary care pathway for patients with inflammatory bowel disease before, during and after pregnancy

**DOI:** 10.1101/2025.02.27.25323005

**Authors:** Els De Dycker, Sien Lenie, Michael Ceulemans, Patricia Geens, Tessy Lambrechts, Elien Loddewijckx, Ariane Paps, Justien Degry, Caroline D’Hondt, Annelies Matthijs, Séverine Vermeire, João Sabino, Bram Verstockt, Lore Lannoo, Kristel Van Calsteren, Marc Ferrante

## Abstract

**Background:** Recent advancements have significantly enhanced our understanding of the interplay between inflammatory bowel disease (IBD) and reproductive health. While international organizations provide guidelines for best practices, translating them into actionable strategies is crucial.

**Aim:** This study aimed to develop a comprehensive care pathway to enhance preconception counselling and support for IBD patients in the perinatal period, ensuring they receive optimal expert care.

**Methods:** We used the 7-phase model for the development of the care pathway.

**Findings:** The resulting care pathway, structured as a time-task matrix, outlines the required actions at preconception, during pregnancy, and in the postpartum period for women with IBD.

**Discussion:** The pathway provides a structured and multidisciplinary approach that addresses the unique needs of IBD patients of childbearing age. It emphasizes holistic and personalized support throughout the preconception, pregnancy, and postpartum period.

**Conclusion:** The development of this care pathway represents a significant advancement in the perinatal management of IBD. By offering multidisciplinary and individualized care, optimal maternal and infant outcomes are pursued, while establishing a new global standard for reproductive health and perinatal management.

**Statement of significance:** *Problem or issue:* Managing IBD in women of childbearing age, particularly those planning pregnancy or already pregnant, is complex and demands a multidisciplinary approach.

*What is already known:* International IBD guidelines outline best practices for managing IBD during pregnancy but implementing these in routine clinical poses significant challenges.

*What this paper adds:* The IBD INFANT Wish care pathway transforms existing guidelines into a structured and multidisciplinary framework, providing comprehensive support and expert care for IBD patients across the preconception, pregnancy, and postpartum period.

## 1. Introduction

Inflammatory bowel diseases (IBD), such as Crohn’s disease and ulcerative colitis, are chronic gastrointestinal conditions requiring long-term multidisciplinary care. These disorders are often diagnosed during the childbearing age (1–3). The variable disease course among IBD patients complicates clinical guidance. While some patients may have prolonged remission, others face continuous relapses, needing intensive medical or surgical treatment. This unpredictable course of the disease significantly impacts patient follow-up in the reproductive period.

When planning a pregnancy, future parents should receive both general and IBD-specific counselling (3). Maintaining women’s health during this period is key, which includes a healthy diet, regular exercise, timely vaccinations, and avoiding substance abuse (2–4). Additionally, women should start folic acid supplements before pregnancy to reduce the risk of neural tube defects. A higher dose of folic acid may be recommended for women taking specific medications such as sulfasalazine, although opinions globally vary (3, 5). Specific IBD-related guidance should address topics as fertility, the impact of IBD and IBD-related treatment on fertility, conception, pregnancy outcomes, breastfeeding and newborn health, but also the impact of pregnancy on IBD severity, and the risk of developing IBD in the offspring.

To optimize pregnancy and infant outcomes, guidelines recommend conceiving during remission and maintaining disease control throughout pregnancy (2, 3, 5, 6). Active IBD at conception or during pregnancy is associated with increased risks of preterm birth, low birth weight, and small for gestational age infants (2, 3, 5–7). Therefore, careful monitoring and appropriate management of disease activity during pregnancy are essential (2, 3, 5). The IBD physician will evaluate whether to adjust or discontinue maintenance therapy, considering gestational age, medical history, and disease activity (2, 4). Mode of delivery should also be part of the discussion, determined on an individual basis and guided by a multidisciplinary team.

In essence, care pathways can significantly enhance the efficiency and quality of care for IBD patients. A care pathway is “a complex intervention for the mutual decision-making and organization of care processes for a well-defined group of patients during a well-defined period” (8). These pathways are developed based on available evidence, including clinical practice guidelines (8, 9). Developing and implementing an “IBD & pregnancy” pathway presents unique challenges due to the unpredictable nature of the disease, particularly before and during pregnancy. A one-size-fits-all approach is not feasible. Some patients may be adequately managed by their local obstetrician, while others will require the expertise of a multidisciplinary team at a tertiary referral centre. Such a team might include an IBD gastroenterologist, obstetrician, colorectal surgeon, IBD dietician, psychologist, and IBD nurse. In this paper, we describe the development and implementation of the IBD Multidisciplinary Family Planning UZ Leuven Care Pathway (IBD INFANT Wish) aimed at improving care quality and efficiency for IBD patients who wish to conceive or are already pregnant. The IBD INFANT Wish pathway seeks to address the complexities of these patients, by offering structured, tailored, multidisciplinary, and evidence-based care.

## 2. Methods

### 2.1 Study design

For the development of the IBD INFANT Wish care pathway at the University Hospitals Leuven, Belgium, we used the 7-phase model developed by the Network of Clinical Paths - KU Leuven. This model guides multidisciplinary teams through the complex process of care pathway development in seven structured phases (8, 10). This manuscript was written using the SQUIRE checklist (see Supplementary Material 1) (11).

#### 2.1.1 Phase 1: Screening phase

The first phase evaluated whether a new pathway would be the optimal solution to achieve the key goals, requiring a thorough understanding of the existing organizational structure and current care process outcomes. The 3-blackboard method, a consensus-building technique, was used (8, 10, 12). Originally developed by the Center for Case Management (Boston, USA) and further refined by the Belgian Dutch Clinical Pathway Network and the European Pathway Association, the approach utilizes three blackboards: the right board outlines the goals of the care pathway, the middle board lists the required activities and interventions to meet the goals, and the left board captures discussion points, bottlenecks, and questions.

#### 2.1.2 Phase 2: Project management phase

This phase was dedicated to defining the care process for the IBD INFANT Wish pathway by means of a group discussion, designed to optimize care for women with IBD who are planning a pregnancy or are already pregnant (8, 10). In addition to agreeing on the definition of the pathway, the IBD INFANT Wish core team and working group were formed.

#### 2.1.3 Phase 3: Diagnostic and objectification phase

The aim of the third phase was to investigate and assess the current organization of the care process from three different perspectives (6, 8): a) the own organization and team, b) patients and their family, and c) the available evidence and legislation.

##### a) The own organization and team

First, the 3-blackboard method was used again to assess the current organizational structure, define objectives, analyse bottlenecks, and identify necessary resources (8, 10, 12). This method ensures a comprehensive, efficient, and goal-aligned pathway development process.

Second, a retrospective study was performed at the University Hospitals Leuven to identify female IBD patients (18-50 years) with an active child wish (upon their hospital visit) who received preconception counselling from IBD physicians and/or obstetricians between January 1, 2017, and December 31, 2023. Patients were identified using the ongoing Crohn’s Disease and Ulcerative Colitis Advanced Research (CCARE) database (S53684), and data were derived from medical records. Ethical approval was obtained from the Ethics Committee Research UZ/KU Leuven (MP022851). Data for the following variables at the time of conception and throughout pregnancy were collected: IBD diagnosis; maternal age at preconception counselling and at conception; presence of perianal Crohn’s disease (active, previous); history of intestinal resection or strictureplasty; IBD-medication (mesalazine, corticosteroids, thiopurines, methotrexate, biologics and/or small molecules) at the time of conception and any change in therapy throughout pregnancy; pregnancy outcomes (miscarriage, stillbirth, mode of delivery, reason for C-section, and intrauterine growth retardation); lifestyle factors (smoking and folic acid intake in pregnancy); and infant outcomes (preterm birth, low birth weight, and small for gestational age).

##### b) The view of IBD patients and their family

To explore the pregnancy-related preferences and childbirth experiences of women with IBD, an online, semi-structured group discussion was organized in collaboration with the Flemish patient organization “Crohn & Colitis Ulcerosa Vereniging – vzw” (CCV-vzw).

##### c) The available evidence and legislation

Recent advancements have greatly enhanced the understanding of the relationship between IBD and reproductive health. The European Crohn’s and Colitis Organization (ECCO) and the American Gastroenterology Association (AGA) have both issued guidelines to inform best practices. In August 2022, ECCO released an updated consensus on sexuality, fertility, pregnancy, and lactation in IBD (5). The AGA’s guidelines align closely with ECCO’s, emphasizing the safety of continuing most IBD therapies during pregnancy and breastfeeding (4). The PIANO Registry (Pregnancy in Inflammatory Bowel Disease and Neonatal Outcomes), a major source of real-world data on the effects of IBD and pharmacological treatments during pregnancy, has contributed to these guidelines (13). These three sources provide robust, evidence-based strategies for managing IBD in the context of reproductive health, underscoring the importance of medication adherence, multidisciplinary care, and patient education to optimize mother-infant outcomes and offer a comprehensive framework for clinicians to support IBD patients with respect to family planning and pregnancy (4, 5, 13).

#### 2.1.4 Phase 4: Development phase

During the fourth phase, a comprehensive care pathway was established, involving a multi-step approach to ensure the pathway’s feasibility, effectiveness, and alignment with clinical guidelines and the care team’s experience (8, 10). Based on the feasibility assessment (see phases 1 to 3), key interventions were created concerning the preconception, pregnancy, and postpartum period. The pathway was finetuned to align with available resources, ensuring interventions were operationally feasible and within the healthcare team’s capabilities. This involved adjusting care plans based on the expertise of gastroenterologists, obstetricians, and other specialists, as well as the availability of support services (8, 10).

#### 2.1.5 Phase 5: Implementation phase

Once the care pathway was established, the implementation phase could start. Implementation involved informing all team members about the care pathway, its use in daily practice and communication to general practitioners (GPs) and obstetricians, both within and outside the hospital.

#### 2.1.6 Phase 6 and 7: Evaluation and follow-up phase

As part of phase 6 of the applied method, the pathway should be evaluated for effectiveness, usability, and compliance. Data collected as part of the evaluation phase could be compared with the baseline results obtained in phase 3 of the development (8, 10). The evaluation (phase 6) and subsequent continuous follow-up (phase 7) are both not within the scope of this paper.

### 2.2 Definitions

The first trimester was characterized as the period of conception until the end of gestational week 13, the second trimester from week 14 until the end of week 27, and the third trimester from week 28 until the delivery (1). Preterm delivery was defined as a birth before 37 weeks of gestation (1). Low birth weight was defined as a birth weight less than 2500 gram, while small for gestational age as birth weight below the 10th percentile with regard to infant sex and gestational age (1, 14), according to the growth charts of the Fetal Medicine Foundation (15). Early miscarriage was defined as an expulsion of a non-viable foetus before the 22nd week of pregnancy; after this period it was called a stillbirth (16). Parity was defined as the number of deliveries of infants aged ≥22 gestational weeks and/or weighing ≥500 gram (16).

### 2.3 Data analysis

Descriptive statistics using IBM SPSS Statistics 29.0.2.0 were applied. For continuous data, medians with interquartile ranges (IQR) were reported, while for discrete data, frequencies and percentages were presented. The discussion with patients was analysed using an inductive, thematic approach.

## 3. Results

### 3.1 Phase 1: Screening phase

The process started with a brainstorming session, leading to a bottleneck analysis, which emphasized the current need for transparency, standardization, effective communication, and improved follow-up care.

### 3.2 Phase 2: Project management phase

The multidisciplinary team brought together stakeholders and experts critical to addressing the complex needs of IBD patients, particularly those with a pregnancy wish or who are currently pregnant (8, 10). The IBD INFANT Wish core team was composed of an IBD gastroenterologist (M.F.), high-risk obstetrician (K.V.C.), and IBD nurse (E.D.D). The working group included IBD gastroenterologists, obstetricians specializing in high-risk pregnancies, colorectal surgeons, IBD nurses, IBD dieticians, case managers in midwifery, and an IBD case manager.

### 3.3 Phase 3: Diagnostic and objectification phase

#### 3.3.1 Care process and project goals

Several care process and project goals were defined. The care process goals were divided according to the preconception, pregnancy, and postpartum period (see Table 1).

**Table 1.** The care process goals for the IBD INFANT Wish care pathway.

| Phase | Care process goals |
| --- | --- |
| Preconception phase | Provide patient education and counselling<br>Health optimization<br>Medication review and adjustment |
| Pregnancy phase | Monitoring and support<br>IBD disease management<br>Assessment and management of pregnancy complications<br>Awareness of specific vaccine limitations for the child<br>Delivery planning |
| Postpartum phase | Postpartum care and support<br>Long-term follow-up |

An overview of the project goals is shown in Table 2. These goals ensure that the pathway is developed, implemented, and continuously improved to achieve the best possible outcomes for the patient and the child.

**Table 2.** The project goals for the IBD INFANT Wish care pathway.

| Project goals | Interpretation |
| --- | --- |
| Establishing evidence-based guidelines | Develop and update evidence-based guidelines for managing pregnant IBD patients based on the latest research and clinical practice |
| Training and education of the team members | Train healthcare providers on specific needs and management strategies for pregnant IBD patients<br>Offer continuous education opportunities to keep healthcare professionals informed about advancements in the field |
| Enhancing patient education and support | Create educational materials and support programs for pregnant IBD patients |
| Quality improvement | Continuously monitor and evaluate the care pathway to identify areas for improvement |
| Interdisciplinary collaboration | Encourage communication and collaboration among healthcare providers (e.g., gastroenterologists, obstetricians, IBD nurses, case managers midwifery, IBD dietitians) |
| Patient-centred care | Tailor care pathways to the individual needs and preferences of patients<br>Engage patients in shared decision-making processes regarding their treatment plans |

#### 3.3.2 Retrospective findings from our hospital database

A review of the CCARE database in our academic hospital identified 908 reproductive female patients. Between January 1, 2017, and December 31, 2023, 52 female IBD patients had a registered, active pregnancy wish and had received preconception counselling from an IBD clinician. In addition, 193 pregnancies were identified among 141 unique individuals, resulting in 158 live births. Of these pregnancies, 57.0% (110/193) received preconception counselling from an IBD clinician. However, more than half of these patients (57.3%; 63/110) had only a notation of an active pregnancy wish in their records, without details on specific counselling by an IBD clinician. For 30.1% of all the pregnancies (58/193), preconception counselling by obstetricians was registered.

The median age at conception was 30 years (IQR: 27.5-33 years). Crohn’s disease was the most prevalent condition (61.7%; n=87). Within the Crohn’s disease group, active and previous perianal involvement was noted for 6 and 14 out of 87 women, respectively. Consequently, 37.6% of patients had ulcerative colitis (n=53) and one patient was known with Inflammatory Bowel Disease type unclassified. In total, 27.9% (n=54) patients had undergone intestinal resection or strictureplasty (the latter only in Crohn’s disease patients).

Maternal characteristics of the pregnant women are displayed in Table 3. Most women (72.4%) had not experienced any abortions. Most women (57.8%) were nulliparous. Among those who had given birth, 31.4% had given birth once. Regarding gravidity, 45.9% of the women were pregnant for the first time, and 30.1% for the second time. Moreover, 12.8% reported smoking during pregnancy, and 99.4% reported using folic acid in pregnancy.

**Table 3.** Maternal characteristics and IBD-related medication use during pregnancy.

| Variables | N | % |
| --- | --- | --- |
| **Maternal characteristics** |  |  |
| Obstetric characteristics (N=185) |  |  |
| Abortion |  |  |
| A0 | 134 | 72.4 |
| A1 | 31 | 16.8 |
| A2 | 15 | 8.1 |
| A3 | 3 | 1.6 |
| A4 | 2 | 1.1 |
| Parity |  |  |
| P0 | 107 | 57.8 |
| P1 | 58 | 31.4 |
| P2 | 15 | 8.1 |
| P3 | 3 | 1.6 |
| P4 | 1 | 0.5 |
| P5 | 1 | 0.5 |
| Gravida |  |  |
| G1 | 85 | 45.9 |
| G2 | 58 | 31.4 |
| G3 | 20 | 10.8 |
| G4 | 12 | 6.5 |
| G5 | 7 | 3.8 |
| G6 | 2 | 1.1 |
| G7 | 0 | 0.0 |
| G8 | 1 | 0.5 |
| Usage during pregnancy |  |  |
| Smoking (N=187) |  |  |
| Yes | 24 | 12.8 |
| No | 163 | 87.2 |
| Folic acid use (N=155) |  |  |
| Yes | 154 | 99.4 |
| No | 1 | 0.6 |
| **Medication use during pregnancy (N=193)** |  |  |
| Mesalazine | 25 | 13.0 |
| Corticosteroids | 5 | 2.6 |
| Thiopurine | 26 | 13.5 |
| Methotrexate | 0 | 0.0 |
| Biological therapy | 126 | 65.3 |
| Infliximab | 40 | 20.6 |
| Adalimumab | 23 | 11.9 |
| Golimumab | 3 | 1.0 |
| Vedolizumab | 34 | 17.6 |
| Ustekinumab | 23 | 11.9 |
| Risankizumab | 2 | 1.0 |
| Small molecules | 0 | 0.0 |
| Medication changed during pregnancy | 84 | 45.1 |
| Biological therapy stopped during pregnancy | 66 | 52.4 |

Results were shown as absolute numbers and percentage. The data relate to unique pregnancies. Individuals can appear in the table with more than one pregnancy. There were 2 multiple pregnancies, resulting in pregnancy outcomes for 195 pregnancies.

With respect to medications, most women (65.3%) used biologics at conception, followed by mesalazine (13.0%) and thiopurines (13.5%). Half (52.4%) of the women stopped their biologic therapy later in pregnancy. No women reported using methotrexate during pregnancy.

Table 4 presents the pregnancy and infant outcomes. Miscarriage occurred in 16.4% of the pregnancies. There were three elective terminations before week 22: one at the patient’s request, one due to extrauterine pregnancy, and one due to genetic abnormalities. Besides, two terminations at week 24 were due to genetic abnormalities and foetal heart defects. Overall, about 40% of the deliveries were C-sections. With respect to adverse infant outcomes, the preterm delivery rate was 8.9%. The median birthweight was 3170 grams (IQR: 2880-3610 grams). Low birth weight and small for gestational age was noted in 7.6% and 12.7% of the infants, respectively. The sample included one set of monozygotic and dizygotic twins.

**Table 4.** Pregnancy and infant outcomes of the IBD pregnancies.

|  | N | % |
| --- | --- | --- |
| **Pregnancy outcome (N=195)** |  |  |
| Live birth | 158 | 81.0 |
| Miscarriage | 32 | 16.4 |
| Elective termination of pregnancy (before week 22) | 3 | 1.5 |
| Pregnancy termination at week 24 | 2 | 1.0 |
| Stillbirth | 0 | 0.0 |
| **Mode of delivery (N=156)** |  |  |
| Vaginal delivery | 93 | 59.6 |
| Caesarean section | 63 | 40.4 |
| Reason of caesarean section |  |  |
| Obstetrical indication | 41 | 65.1 |
| Patient with perianal fistulising disease | 14 | 22.2 |
| Patient with a pouch | 5 | 7.9 |
| Patient's wish | 1 | 1.6 |
| Other | 2 | 3.2 |
| **Infant outcomes (N=158)** |  |  |
| Preterm delivery<sup>a</sup> | 14 | 8.9 |
| Low birth weight<sup>b</sup> | 12 | 7.6 |
| Small for gestational age<sup>c</sup> | 20 | 12.7 |
| Intra-uterine growth retardation (N=143) | 9 | 6.3 |

Results were shown as absolute numbers and percentage. The data relate to unique pregnancies. There were 2 multiple pregnancies, resulting in pregnancy outcomes for 195 pregnancies. Individuals can appear in the table with more than one pregnancy. <sup>a</sup>preterm delivery is defined as a delivery before 37 weeks of pregnancy; <sup>b</sup>low birth weight is defined as a birth weight less than 2500 gram; <sup>c</sup>small for gestational age is defined as birth weight below the 10th percentile according to gestational age and infant sex.

#### 3.3.3 The view of IBD patients

The focus group consisted of 6 women of reproductive age, each bringing unique experiences with pregnancy and family planning. Five women had given birth to one or more children, while one women was actively trying to conceive but had not been pregnant yet. Among participants, three were diagnosed with ulcerative colitis, including the woman who had not yet conceived, and three had Crohn’s disease. Both vaginal and caesarean deliveries were represented among the participants. One participant had undergone pouch surgery before pregnancy, and another had a permanent ileostomy during her pregnancy.

One key finding was the importance of multidisciplinary care. Women emphasized the benefits of having a coordinated team of healthcare providers, including gastroenterologists, obstetricians with IBD knowledge, IBD nurses, and IBD dietitians, who could work together to tailor a care plan addressing both their IBD and pregnancy needs. Women also acknowledged the significance of emotional support, as managing IBD during pregnancy can be emotionally challenging due to unpredictability and complexity.

Another significant finding was the prohibition of administering live attenuated vaccines to infants exposed to biological agents in utero. Several women noted that their medication use during pregnancy was often not questioned by the physician at “Kind en Gezin,” which could have led to the administration of live attenuated vaccines if they had not proactively mentioned their medication history. The discussions also highlighted the need for information on how prior colorectal surgery could impact fertility, pregnancy, and

### 3.4 Phase 4: Development phase

#### 3.4.1 Development of the care pathway IBD INFANT Wish

Upon completion of the first three phases, we developed the IBD INFANT Wish care pathway. Different patient trajectories, expected risks, and comorbidities were considered, ensuring that each patient’s unique circumstances were addressed. The first step was creating a decision tree detailing how to manage IBD patients and their need for scheduling preconception advice. This depended on how they described their desire for children (none, latent, or short-term) or if they were already pregnant, and determines if the patient needs preconception and/or perinatal counselling from both the IBD physician and/or a (high-risk) pregnancy obstetrician. The care pathway, as shown in Figure 1, is structured as a time-task matrix. It outlines the necessary actions for a female IBD patient from active pregnancy desire to the postpartum phase.

**Figure 1.**
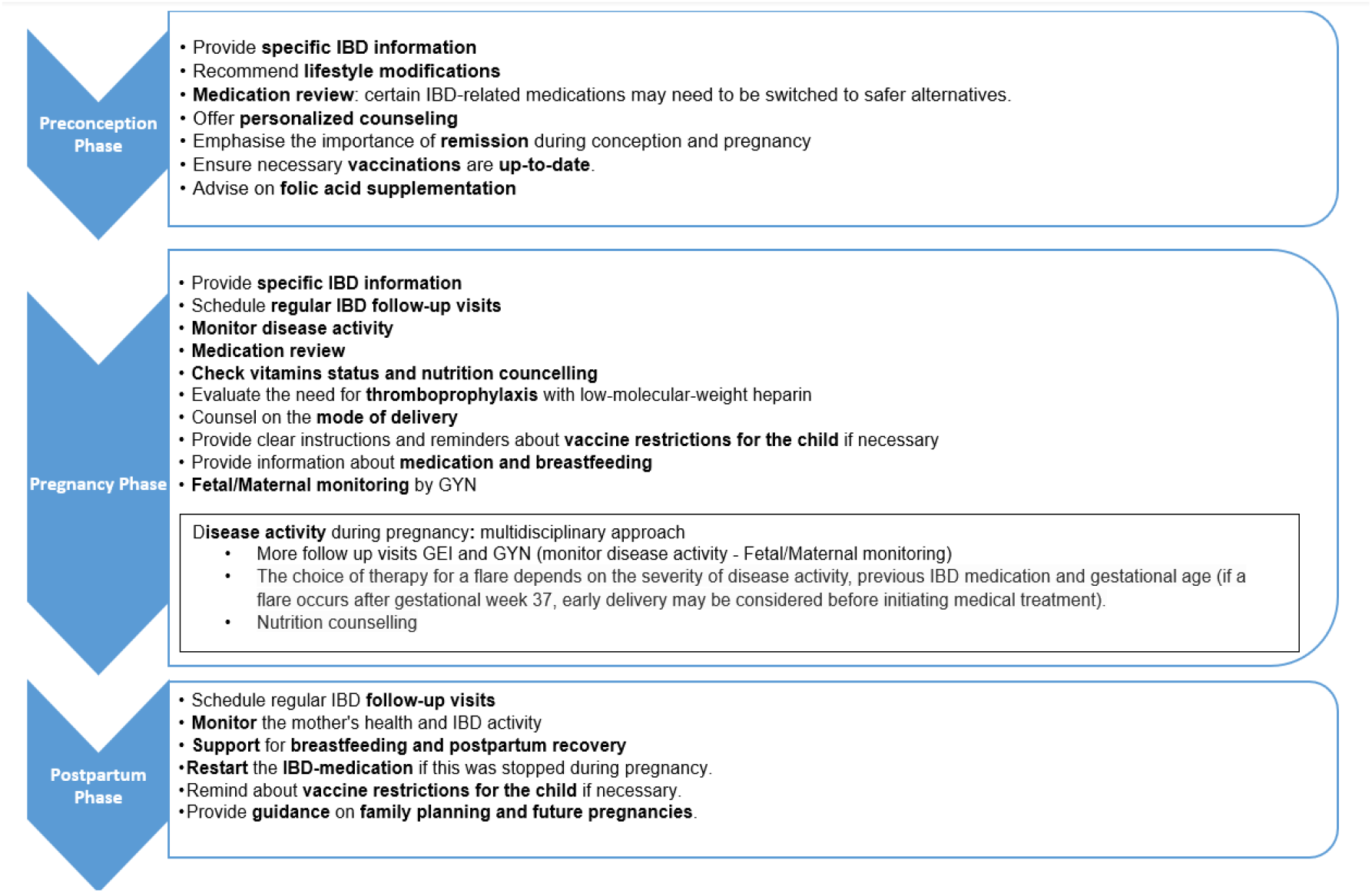
Time - task matrix of the IBD INFANT Wish care pathway.

##### Preconception phase

Providing tailored information to IBD patients is crucial and should cover topics such as fertility, heredity, the impact of IBD and related medications on fertility and conception, the effects of pregnancy on IBD, and delivery methods. IBD patients may choose to remain childless since 3-5% will inherit the disease if one parent has IBD, increasing to 30% if both parents are affected (5).

Women with IBD have similar fertility rates to healthy women, if the disease is in remission and there is no history of lower abdominal surgery. Laparoscopic surgery may reduce infertility risks (5). It is essential to discuss how active IBD can decrease fertility and emphasize that achieving clinical remission can significantly enhance conception chances (5).

A thorough medication review prior to conception is necessary, as certain IBD medications, such as methotrexate, Janus kinase inhibitors, and spingosine-1-phosphate receptor modulators, are contraindicated in pregnancy (5). Personalized counselling should address specific patient concerns and fears and provide targeted advice based on individual circumstances. Achieving and maintaining remission at the time of conception and during pregnancy, ideally confirmed by endoscopy, intestinal ultrasound and/or the biomarker faecal calprotectin, is crucial for both mother and (unborn) infant.

##### Pregnancy phase

When a patient informs the IBD team of an ongoing pregnancy, the frequency of follow-up visits should be increased. Typically, this includes one IBD outpatient clinic visit per trimester for patients in remission (especially for patients under advanced therapy) to closely monitor the mother’s condition and ensure sustained remission. More frequent visits are recommended in patients with active disease. Additionally, regular appointments with an obstetrician should be scheduled for maternal-foetal monitoring. For patients who have not received preconception advice, it is essential to provide comprehensive information about IBD, its potential impact on pregnancy, associated risks, and necessary precautions. Faecal calprotectin should be tested once per trimester during remission, as it is a reliable marker for assessing disease activity in pregnancy. While certain blood parameters, such as haemoglobin and CRP, may be less reliable due to physiological changes in pregnancy, observing their trends can still yield valuable insights (5). Additionally, intestinal ultrasound can be an effective and accurate tool for monitoring disease activity in pregnant IBD patients (5).

A thorough review of medication is also important at this stage. For women with active disease at conception or during pregnancy, or with difficult-to-control disease, continuing biologics throughout pregnancy is strongly recommended (5). Also, for women in remission, discontinuing therapy before the third trimester is generally discouraged, as it can increase the risk of relapse and result in unfavourable pregnancy outcomes. However, if a patient in long-term remission wishes to stop anti-TNF therapy before the third trimester, resuming treatment soon after delivery is recommended (5). For women in remission treated with non-TNF biologics, such as ustekinumab or vedolizumab, decisions on discontinuation should be individualized, weighing the risk of relapse against the limited data on foetal exposure (5). At our institution, the continued use of vedolizumab, ustekinumab, and newer anti-IL23 inhibitors is supported throughout pregnancy. It is also important to educate parents about vaccination restrictions for the child. Infants exposed to biologics in utero should not receive live attenuated vaccines during the first year of life or until the biologic is no longer detectable in the infant’s blood (5). To ensure healthcare providers are informed, a sticker notification can be placed in the child’s health booklet (provided by the perinatal organization Kind & Gezin), indicating the mother’s use of biological therapy during pregnancy.

Medications considered low-risk in pregnancy are also regarded as low-risk during breast-feeding and can typically be continued (5). Providing clear, detailed information on the safety of these medications during breastfeeding is essential to support informed decision-making.

Pregnant women with IBD require special attention to their nutritional needs. Adequate weight gain during pregnancy is important, as insufficient weight gain is associated with adverse infant outcomes (5). If weight gain is inadequate, tailored nutritional advice will be provided by IBD dieticians to address this concern. With regard to method of delivery, vaginal delivery is generally preferred. However, a caesarean section is recommended in case of both prior and active perianal and rectovaginal fistulising disease. For patients with an ileal pouch anal anastomosis, both a caesarean section and vaginal delivery can be considered (2, 3, 5)

In the event of an IBD flare during pregnancy, a multidisciplinary approach is essential (5). The choice of pharmacological treatment should be guided by the severity of the disease, previous IBD medication use, and the gestational age (5). If a flare occurs after 37 weeks of gestation, early delivery may be considered before initiating medical treatment (5). Comprehensive management should include nutritional counselling to ensure both maternal and foetal health, alongside regular monitoring of disease activity. Continuous maternal and foetal monitoring is critical for promptly addressing complications and adjusting treatment plans as needed.

Both pregnancy, caesarean delivery, and active IBD increase the risk of venous thromboembolism. Therefore, evaluating the overall thromboembolic risk during pregnancy and postpartum is important, and when significantly increased, low-molecular weight heparins should be recommended (RCOG guideline) (5).

##### Postpartum phase

An increased risk of postpartum flares, linked to stress, sleep deprivation or alterations in the immune system, exists. If biological therapy was stopped during pregnancy, it is vital to restart it postpartum to maintain remission, prevent flare-ups, and ensure stable maternal health. It is also essential to confirm whether the patient is breastfeeding, as medications deemed low risk during pregnancy are also safe during breastfeeding and can be resumed (5). Postpartum care for IBD patients should include guidance on family planning and future pregnancies. This involves discussing the optimal timing for subsequent pregnancies, potential risks, and strategies to maintain disease control.

#### 3.4.2 Development of materials

To address the unique needs of IBD patients, we developed five different information materials.

First, we created an information banner to be displayed at the infusion unit and outpatient clinic TV screens, highlighting the importance of preconception care. This banner aims to educate IBD patients on optimizing maternal health, preventing pregnancy complications, and the importance of medication review and lifestyle adjustments, encouraging them to seek preconception advice.

Second, we developed a letter template to inform GPs and peripheral obstetricians about the preconception counselling their IBD patient received. The template includes recommendations on disease and medication management, delivery methods, and other critical aspects of preconception care for this specific patient population.

Thirdly, we designed a comprehensive “Inflammatory Bowel Disease and Pregnancy” brochure for IBD patients wishing to conceive or who are already pregnant. This brochure provides both general and IBD-specific preconception information and can be used by IBD and obstetric teams. It is informative but does not replace individualized preconception consultations.

Fourthly, a detailed decision tree for scheduling preconception advice was developed. This tool will help the IBD team to deliver optimal care to patients with a pregnancy wish or who are already pregnant, with special attention to high-risk pregnancies due to factors such as complicated pregnancy history, repeated miscarriages, preterm birth, psychiatric disorders, and other immune-mediated inflammatory disorders.

Ultimately, and equally important, for female IBD patients who received biologic therapy during pregnancy, a sticker notification was created to be placed in the child’s “Kind en Gezin” booklet. This informs healthcare professionals that the mother received biologic therapy in pregnancy. Currently, the vaccination schedules of the perinatal organizations in Belgium include the live attenuated rotavirus vaccine to be administered before week 24 of life. Other live attenuated vaccines that should be avoided include the yellow fever vaccine and the Bacillus Calmette-Guérin (BCG) vaccine, although these are not part of the basic vaccination scheme for infants in their first year of life in Belgium (5).

## 4. Discussion

Conception and pregnancy are key life events, and for women of reproductive age with IBD, these moments can be accompanied by heightened fear and anxiety. To optimize maternal and child health in future IBD pregnancies and strengthen multidisciplinary collaboration, we developed the practical IBD INFANT Wish care pathway for patients with a pregnancy wish or already pregnant. The development process was guided by the 7-phase model of the Network of Clinical Paths KU Leuven, which emphasizes setting achievable goals (8). The involvement of a multidisciplinary team, including IBD gastroenterologists, high-risk obstetricians, colorectal surgeons, IBD nurses, dieticians, midwifery case managers, and an IBD case manager, was integral to the pathway’s success, underscoring its value and potential.

The ECCO guidelines, AGA recommendations, and PIANO registry results served as essential documents for designing the pathway. These evidence-based resources emphasize key strategies such as medication adherence, multidisciplinary care, and patient education to improve outcomes for both mothers and their (unborn) children. Such resources provide a comprehensive framework for healthcare professionals to support IBD patients during family planning, pregnancy, and beyond (4, 5, 13). To ensure a patient-centred approach, we also gathered insights from IBD patients and the IBD patient organization, avoiding that patient-related needs or preferences were overlooked.

Following the implementation of the IBD INFANT Wish care pathway in our academic hospital, its effectiveness, usability, and compliance must be systematically evaluated. This process should include collecting feedback from patients and healthcare providers, analysing clinical outcomes, and measuring patient satisfaction. Materials developed for the pathway, such as banners, brochures, and decision trees, should also be regularly reviewed and updated. The retrospective medical records’ data collected during phase 3 of the development provide baseline measurements for mother-child outcomes in IBD pregnancies at our institution. These data will be essential for future evaluations of the pathway’s effectiveness. We plan to conduct a comparative analysis one year after implementation, investigating trends in medication use or switching around conception and during pregnancy, as well as mother-infant outcomes. The goal is to sustain and further enhance the care pathway, ensuring it leads to high-quality, evidence-based care for IBD patients planning or experiencing pregnancy. Our care pathway may also be instructive for our international institutions considering the implementation of a structured approach for the perinatal management of IBD patients.

Previous research has demonstrated that active IBD is associated with higher rates of low birth weight, preterm birth, small for gestational age infants, miscarriage, and stillbirths (2, 3, 6). In our retrospective analysis from a tertiary setting, the relatively low rates of low birth weight (7.9% vs. 6.6%) and preterm delivery (8.9% vs. 8.1%), in comparison to overall perinatal statistics in Flanders, Belgium, are encouraging but should be cautiously interpreted (17). The high C-section rate in our cohort (40%) may reflect the complex clinical considerations in managing IBD pregnancies but should be further investigated.

With regard to some methodological considerations, the care pathway was developed from the perspective of a tertiary, university hospital. The baseline measurement of IBD pregnancies is limited in terms of numbers as it relies on the medical records from a single centre. By its nature, our data carry the risk of missing outcome information, particularly for (healthier) cases where deliveries occurred in other regional hospitals. To gain insight into perinatal experiences of IBD patients, we organized a single group discussion, identifying some key challenges faced by patients. While individual interviews could have provided a more detailed understanding of these experiences, this approach was not feasible due to practical constraints. Interviews should be considered in the future to ensure a more comprehensive exploration of patient experiences. Additionally, including a paediatrician and patient representative in our working group could enhance future discussions by bringing in more diverse perspectives (e.g., with respect to the vaccination of infants). Finally, no objective data have been collected yet on the perspectives of GPs, peripheral obstetricians and the perinatal organizations in Belgium regarding the new care pathway, warranting further exploration in the future.

## 5. Conclusion

The development and implementation of the IBD INFANT Wish care pathway at our institution represents a significant advancement in the perinatal management of female IBD patients. This pathway combines a multidisciplinary approach with evidence-based practices to optimize care, improve maternal-infant outcomes, and provide comprehensive and consistent support and information in the perinatal period. It ensures that patients receive tailored, patient-centred care that addresses both their IBD and pregnancy-related needs. This pathway sets a global benchmark for managing preconception and pregnancy care in female IBD patients. However, continuous evaluation following its implementation, along with subsequent modifications and updates, will be essential to maintain and maximize its relevance and effectiveness.

## Data Availability

All data produced in the present work are contained in the manuscript.

## Declaration of Generative AI and AI-assisted technologies in the writing process

During the preparation of this work, the authors used Copilot in order to enhance the quality of English in the manuscript. After using this tool, the authors reviewed and edited the content as needed and take full responsibility for the content of the publication.

## Disclaimer

The terms “women,” “woman,” “female” and “maternal” are used in this manuscript to refer to all those who are (almost) pregnant and give birth. The authors acknowledge that not all people who are (almost) pregnant and give birth will identify themselves as women.

## CRediT authorship contribution statement

Els De Dycker: Conceptualization, Data curation, Formal analysis, Investigation, Methodology, Project administration, Software and Writing – original draft. Sien Lenie: Writing – original draft. Michael Ceulemans: Writing – original draft. Patricia Geens: Reviewing manuscript. Tessy Lambrechts: Reviewing manuscript. Elien Loddewijckx: Reviewing manuscript. Ariane Paps: Reviewing manuscript. Justien Degry: Identifying patient data, Reviewing manuscript. Caroline D’Hondt: Reviewing manuscript. Annelies Matthijs: Reviewing manuscript. Séverine Vermeire: Reviewing manuscript. João Sabino: Reviewing manuscript. Bram Verstockt: Reviewing manuscript. Lore Lannoo: Conceptualization and Reviewing manuscript. Kristel Van Calsteren: Conceptualization and Reviewing manuscript. Marc Ferrante: Conceptualization, Investigation, Methodology, Supervision and Reviewing manuscript.

## Conflicts of interest

E.D.D. reports no conflicts of interest; S.L. reports no conflicts of interest; M.C. is coordinator of the BELpREG pregnancy registry in Belgium which received research grants from P&G Health, Tilman, UCB, Almirall, KelaPharma, Sanofi, and Johnson & Johnson. The companies providing research grants have no role in the design of the study; in collection, analysis, or interpretation of data; in the writing of the manuscript, or in the decision to publish the results. M.C. received speaker’s fees from UCB and KelaPharma, and a research grant from HRA Pharma; P.G. reports no conflicts of interest; T.L. reports no conflicts of interest; E.L. reports no conflicts of interest; A.P. reports no conflicts of interest; J.D. reports no conflicts of interest; C.D’H. reports no conflicts of interest; A.M. reports no conflicts of interest; S.V. received lecture fees from AbbVie, Dr. Falk Pharma, Ferring, Hospira, MSD, Takeda and Tillotts; consultancy fees from AbbVie, AbolerIS Pharma, Alimentiv, Arena, AstraZeneca, Avaxia, BMS, Boehringer Ingelheim, Celgene, CVasThera, Dr Falk Pharma, Eli Lilly, Ferring, Galapagos, Genentech/Roche, Gilead, Hospira, Imidomics, Janssen, Johnson and Johnson, Materia Prima, MiroBio, Morphic, MrMHealth, MSD, Mundipharma, Pfizer Inc, Prodigest, Progenity, Prometheus, Robarts Clinical Trials, Second Genome, Shire, Surrozen, Takeda, Theravance Biopharma, Tillots Pharma AG and Zealand Pharma; and grant/research support from AbbVie, Galapagos, MSD, Pfizer Inc and Takeda; J.S. received speaker’s fees from Pfizer, Abbvie, Ferring, Falk, Takeda, Janssen, Fresenius, and Galapagos; Consultancy fees from Pfizer, Janssen, Ferring, Fresenius, Abbvie, Galapagos, Celltrion, Pharmacosmos, and Pharmanovia; Research support from Galapagos and Viatris; B.V. received research support from AbbVie, Biora Therapeutics, Celltrion, Landos, Pfizer, Sossei Heptares, and Takeda. Additionally, B.V. received speaker’s fees from AbbVie, Biogen, Bristol Myers Squibb, Celltrion, Chiesi, Eli Lilly, Falk, Ferring, Galapagos, Johnson and Johnson, MSD, Pfizer, R-Biopharm, Sandoz, Takeda, Tillots Pharma, Truvion, and Viatris. Furthermore, B.V. received consultancy fees from AbbVie, Alfasigma, Alimentiv, Applied Strategic, AstraZeneca, Atheneum, BenevolentAI, Biora Therapeutics, Boxer Capital, Bristol Myers Squibb, Eli Lilly, Galapagos, Guidepont, Landos, Merck, Mylan, Nxera, Inotrem, Ipsos, Johnson and Johnson, Pfizer, Progenity, Sandoz, Sanofi, Santa Ana Bio, Sapphire Therapeutics, Sosei Heptares, Takeda, Tillots Pharma, and Viatris. Lastly, B.V. holds stock options in Vagustim. The companies providing research grants have no role in the design of the study; in collection, analysis, or interpretation of data; in the writing of the manuscript, or in the decision to publish the results; L.L. reports no conflicts of interest; K.V.C. reports no conflicts of interest;M.F. received research grants from AbbVie, EG Pharma, Janssen, Pfizer, Takeda and Viatris; consultancy fees from AbbVie, AgomAb Therapeutics, Boehringer Ingelheim, Celgene, Celltrion, Eli Lilly, Janssen-Cilag, Merck Sharp and Dohme, MRM Health, Pfizer, Takeda and ThermoFisher; and speakers’ fees from AbbVie, Biogen, Boehringer Ingelheim, Dr Falk Pharma, Ferring, Janssen-Cilag, Merck Sharp and Dohme, Pfizer, Takeda, Truvion Healthcare and Viatris.

## Acknowledgements and disclosures

This manuscript is based on the master thesis Nurse Specialist of Els De Dycker at KU Leuven, Leuven, Belgium. J.S. and M.F. are senior clinical investigators of the Research Foundation Flanders (FWO). B.V. is supported by the Clinical Research Fund (KOOR) at UZ Leuven and the Research Council at KU Leuven. S.V. holds a BOF-KFO from the KU Leuven. M.C. is supported by a Senior Postdoctoral Fellowship fundamental research of the Research Foundation Flanders (FWO, 1246425N).

## Notes

### Competing Interest Statement

EDD reports no conflicts of interest. SL reports no conflicts of interest. MC is coordinator of the BELpREG pregnancy registry in Belgium which received research grants from P&G Health, Tilman, UCB, Almirall, KelaPharma, Sanofi, and Johnson & Johnson. The companies providing research grants have no role in the design of the study; in collection, analysis, or interpretation of data; in the writing of the manuscript, or in the decision to publish the results. M.C. received speakers fees from UCB and KelaPharma, and a research grant from HRA Pharma; P.G. reports no conflicts of interest; T.L. reports no conflicts of interest; E.L. reports no conflicts of interest; A.P. reports no conflicts of interest. JD reports no conflicts of interest. CDH reports no conflicts of interest; A.M. reports no conflicts of interest; S.V. received lecture fees from AbbVie, Dr. Falk Pharma, Ferring, Hospira, MSD, Takeda and Tillotts; consultancy fees from AbbVie, AbolerIS Pharma, Alimentiv, Arena, AstraZeneca, Avaxia, BMS, Boehringer Ingelheim, Celgene, CVasThera, Dr Falk Pharma, Eli Lilly, Ferring, Galapagos, Genentech/Roche, Gilead, Hospira, Imidomics, Janssen, Johnson and Johnson, Materia Prima, MiroBio, Morphic, MrMHealth, MSD, Mundipharma, Pfizer Inc, Prodigest, Progenity, Prometheus, Robarts Clinical Trials, Second Genome, Shire, Surrozen, Takeda, Theravance Biopharma, Tillots Pharma AG and Zealand Pharma; and grant/research support from AbbVie, Galapagos, MSD, Pfizer Inc and Takeda; JS received speakers fees from Pfizer, Abbvie, Ferring, Falk, Takeda, Janssen, Fresenius, and Galapagos; Consultancy fees from Pfizer, Janssen, Ferring, Fresenius, Abbvie, Galapagos, Celltrion, Pharmacosmos, and Pharmanovia; Research support from Galapagos and Viatris; B.V. received research support from AbbVie, Biora Therapeutics, Celltrion, Landos, Pfizer, Sossei Heptares, and Takeda. Additionally, B.V. received speaker's fees from AbbVie, Biogen, Bristol Myers Squibb, Celltrion, Chiesi, Eli Lilly, Falk, Ferring, Galapagos, Johnson and Johnson, MSD, Pfizer, R-Biopharm, Sandoz, Takeda, Tillots Pharma, Truvion, and Viatris. Furthermore, B.V. received consultancy fees from AbbVie, Alfasigma, Alimentiv, Applied Strategic, AstraZeneca, Atheneum, BenevolentAI, Biora Therapeutics, Boxer Capital, Bristol Myers Squibb, Eli Lilly, Galapagos, Guidepont, Landos, Merck, Mylan, Nxera, Inotrem, Ipsos, Johnson and Johnson, Pfizer, Progenity, Sandoz, Sanofi, Santa Ana Bio, Sapphire Therapeutics, Sosei Heptares, Takeda, Tillots Pharma, and Viatris. Lastly, B.V. holds stock options in Vagustim. The companies providing research grants have no role in the design of the study; in collection, analysis, or interpretation of data; in the writing of the manuscript, or in the decision to publish the results. LL reports no conflicts of interest. KVC reports no conflicts of interest. MF received research grants from AbbVie, EG Pharma, Janssen, Pfizer, Takeda and Viatris; consultancy fees from AbbVie, AgomAb Therapeutics, Boehringer Ingelheim, Celgene, Celltrion, Eli Lilly, Janssen-Cilag, Merck Sharp and Dohme, MRM Health, Pfizer, Takeda and ThermoFisher; and speakers fees from AbbVie, Biogen, Boehringer Ingelheim, Dr Falk Pharma, Ferring, Janssen-Cilag, Merck Sharp and Dohme, Pfizer, Takeda, Truvion Healthcare and Viatris.

### Funding Statement

This study did not receive any funding.

### Author Declarations

Ethics Committee Research of the University Hospitals / KU Leuven university in Leuven, Belgium gave ethical approval for this work (S53684 - MP022851).

